# Real World Fertility Evaluation & Care Prior to In Vitro Fertilization: Care Gaps That Could be Addressed by Restorative Reproductive Medicine

**DOI:** 10.64898/2026.07.13.26357941

**Authors:** Tracey Parnell, Monica Minjeur, Craig Turczynski, Tony Pistilli

**Affiliations:** International Institute for Restorative Reproductive Medicine, Crawley, West Sussex, UK; Department of Family Medicine, University of British Columbia, Vancouver, BC, Canada; Radiant Clinic, Cedar Rapids, IA, USA; RRM Diagnostics, Dallas, TX, USA; Duquesne University College of Osteopathic Medicine, Pittsburgh, PA, USA; Axene Health Partners, McKinney, Texas, USA

**Keywords:** infertility, in vitro fertilization, IVF, ASRM, AUA/ASRM, guideline adherence, reproductive endocrinology, restorative reproductive medicine, RRM, diagnosis-directed care, claims analysis

## Abstract

**Objective:** To evaluate adherence to published American Society for Reproductive Medicine (ASRM) and American Urological Association (AUA) infertility evaluation and treatment recommendations among commercially insured infertility patients who subsequently underwent in vitro fertilization (IVF), and to assess whether observed care gaps support the need for a restorative reproductive medical framework.

**Methods:** A retrospective claims-based analysis was performed using MarketScan® Commercial Claims and Encounter Data between January 1, 2021, and December 31, 2024. Approximately five million commercially insured members were evaluated. Patients with infertility-related diagnoses who subsequently underwent IVF were identified. Claims were analyzed for evidence of selected diagnostic testing, medical treatment, or surgical interventions recommended by ASRM or AUA/ASRM guidance for specific infertility-related diagnoses before IVF initiation. Cumulative adherence rates were assessed up to nine months following initial infertility diagnosis and compared with the timing of initiating IVF.

**Results:** IVF initiation started early and consistently preceded completion of most recommended evaluations and treatments. By 3 months, the proportion of patients with IVF initiation ranged from 28% to 39% across different infertility-related diagnoses, while adherence to most recommended interventions remained low. Overall, by 9 months, IVF utilization had reached 70–85%, while many recommended evaluations and treatments remained below 40% adherence, with several interventions remaining below 15%. Observed care gaps (percentage of patients receiving IVF prior to other evaluation or treatment recommendations) ranged from approximately 13% to 78% for most recommended evaluations and treatments, with several measures demonstrating gaps over 50% of patients.

**Discussion:** These findings suggest substantial divergence between published infertility-care recommendations and observed practice patterns. From the perspective of restorative reproductive medicine (RRM), the gaps are clinically important because many recommended steps are directed toward identifying, correcting, restoring, or preserving reproductive function and anatomy before offering or initiating IVF treatment. Limitations to these data include lack of individualized patient medical data, preferences, or circumstances, and not capturing any treatment that was not reimbursed by commercial insurance (i.e., any cash payment for services). In addition, the recommendations assessed have variable levels of underlying evidence and likely vary in the level of their acceptance among practicing clinicians.

**Conclusions:** Many commercially insured infertility patients progressed to IVF without documented evidence of diagnostic evaluation or therapeutic interventions recommended in ASRM and AUA/ASRM guidance. These findings raise important questions regarding the implementation of adequate evaluation before IVF and the extent to which patients receive meaningful opportunities for diagnosis-directed treatment of potentially reversible causes of infertility. The findings further suggest an important role for RRM as a quality-of-care framework focused on comprehensive evaluation, correction of underlying dysfunction, preservation of reproductive anatomy and physiology, and optimization of patient-centered fertility care, prior to considering IVF.

## INTRODUCTION

Infertility may be approached through fundamentally different clinical pathways. Restorative reproductive medicine (RRM) seeks to identify, correct, restore, and preserve reproductive function and anatomy wherever possible.^1^ This approach requires a thorough diagnostic evaluation, recognition of underlying reproductive, endocrine, metabolic, inflammatory, immunologic, anatomic, genetic and gamete specific factors for both male and female patients, and corrected treatment targeted to the identified factors, with sufficient time for condition-specific treatment to restore reproductive function. In contrast, in vitro fertilization (IVF) can achieve pregnancy more rapidly by bypassing aspects of natural reproductive function through extracorporeal fertilization and embryo transfer.^2^ In the United States, assisted reproductive technologies (ART), particularly in IVF, have become increasingly utilized as treatment for infertility, with the number of IVF cycles more than doubling over the past decade and approximately 2% of U.S. births now resulting from IVF.^3^ While IVF may provide an effective pathway to pregnancy for many patients, IVF may easily be undertaken without fully addressing underlying causes of infertility.

This distinction is clinically relevant, in part because reproductive endocrinology and infertility care, even when IVF is ultimately used, is still expected to include diagnostic evaluation and treatment of known or suspected contributors to infertility. Reproductive professional society guidelines do not describe IVF as a substitute for evaluation of ovulatory function, tubal and uterine anatomy, endocrine disorders, endometriosis, male-factor infertility, or other potentially reversible conditions. Rather, assisted reproductive technologies are generally situated within a broader framework of treatment options following infertility evaluation and condition-specific management.

The American Society for Reproductive Medicine (ASRM), for example, states that “the infertility evaluation should include an evaluation of ovulatory status, the structure and patency of the female reproductive tract, and semen evaluation of the male partner” (p. 1257).^4^ ASRM guidance further emphasizes identification and management of potentially reversible reproductive, endocrine, metabolic, and anatomical contributors to infertility. Recommended elements of care include evaluation and treatment of endocrine disorders such as hypothyroidism and hyperprolactinemia, comprehensive diagnostic evaluation of amenorrhea, ovulation induction for anovulatory infertility associated with polycystic ovary syndrome, consideration of expectant management or superovulation with intrauterine insemination for selected patients with endometriosis-associated infertility, and surgical management of conditions such as endometriosis when clinically indicated.^5–7^

Similarly, the jointly authored American Urological Association/American Society for Reproductive Medicine male infertility guidelines emphasize comprehensive male-factor evaluation and identification of potentially reversible causes of infertility. These recommendations include repeated semen analysis, endocrine evaluation with hormonal testing in selected populations, genetic and anatomic evaluation for men with azoospermia or severe oligospermia, assessment of sperm DNA integrity and karyotype in selected recurrent pregnancy loss populations, and treatment of clinically significant conditions such as varicocele when appropriate.^8–9^ Collectively, these recommendations affirm that infertility care should include evaluation of both partners and treatment of identifiable contributors when clinically indicated, prior to considering or pursuing ART.

RRM places these and other diagnostic and therapeutic steps at the center of infertility care.^10^ Its clinical priority is not merely to achieve pregnancy, but to restore reproductive health, correct dysfunction, preserve reproductive anatomy and physiology, and support natural conception when feasible.^11^ From an RRM perspective, an important clinical and policy question is whether patients are being given a meaningful opportunity for diagnosis-directed treatment before proceeding to IVF, consistent with professional guidelines that recommend evaluation and treatment of known contributors to infertility.

Patient preferences research suggests that many individuals experiencing infertility desire a more comprehensive diagnostic evaluation before proceeding to assisted reproductive technologies such as in vitro fertilization (IVF).^12^ In a secondary analysis of two nationally representative U.S. surveys, 70% of respondents indicated a preference for fertility care that identifies and treats underlying causes of infertility.^13^ Of respondents who received IVF, 83% expressed significant concern about undiagnosed health conditions 46% of them were ‘very concerned’. This raises the possibility that couples may be directed toward IVF before potentially contributing factors have been fully evaluated.

However, the ASRM has argued that standard infertility care includes “thorough diagnostic workups” and “treatment of underlying conditions” and that it is a “false narrative” to suggest “that standard fertility care skips proper diagnosis and healing.”^14^ These statements raise an empirical question: how consistently are guideline-recommended diagnostic and therapeutic steps completed before IVF in real-world practice? If standard infertility care routinely identifies and treats reversible or contributing conditions before IVF, then claims-based practice patterns should demonstrate high adherence to relevant ASRM and AUA/ASRM recommendations. If, however, substantial proportions of patients proceed to IVF without documented evidence of recommended evaluation or treatment, this would suggest a gap between professional-society standards and observed clinical practice.

Administrative claims databases provide an opportunity to evaluate real-world infertility-care pathways across large commercially insured populations.^15–17^ Such analyses can help determine whether guideline-recommended diagnostic and therapeutic steps are completed before IVF initiation and whether contemporary infertility practice provides the diagnostic and restorative opportunity that many patients prefer and that professional guidelines recommend.

The purpose of this study was to evaluate adherence to selected ASRM and AUA/ASRM infertility recommendations among commercially insured patients who ultimately proceeded to IVF. By examining documented care prior to IVF initiation, this study assesses whether real-world IVF pathways reflect guideline-concordant evaluation and treatment of underlying reproductive dysfunction before proceeding to IVF.

## METHODS

### Study Design

This study utilized a retrospective observational design based on administrative claims data.

### Data Source

The analysis was conducted using the MarketScan® Commercial Claims and Encounter Database covering the period from January 1, 2021, through December 31, 2024. The database contains adjudicated inpatient, outpatient, professional, and retail pharmacy claims from a large commercially insured population. Approximately five million covered members were available for analysis.

Longitudinal member tracking and family linkage capabilities allowed evaluation of infertility-related diagnostic and treatment patterns over time. IVF and intrauterine insemination (IUI) treatment episodes were identified using ICD-10-CM diagnosis codes, CPT and HCPCS procedure codes, and NDC-based pharmacy therapeutic classes.

### Cohort Identification & Guideline Adherence

Cohorts were constructed by identifying patients with infertility-related ICD-10 diagnosis codes and then restricting the analytic sample to those who subsequently underwent in vitro fertilization during the 15-month observation period following the index date. The index date was defined as the first qualifying infertility diagnosis, and patients were followed longitudinally from that point to assess whether guideline-recommended evaluation or treatment occurred before IVF initiation.

Male cohorts were defined according to diagnoses with corresponding recommendations in the AUA/ASRM male infertility guideline and included general male infertility, varicocele-associated infertility, azoospermia, and recurrent pregnancy loss.

Female cohorts were defined using infertility-related diagnoses that aligned with clinically operationalizable recommendations from ASRM guidance documents.^4-7,18^ These included polycystic ovary syndrome, hyperprolactinemia, endometriosis-associated infertility, dysmenorrhea as a proxy for suspected endometriosis, hypothyroidism, amenorrhea, oligomenorrhea, anovulation, and/or androgen excess.

We selected diagnostic evaluations, medical therapies, and surgical interventions, which are recommended in ASRM and AUA/ASRM guidance for patients with specific underlying diagnoses, and which provide sufficiently direct recommendations to support consistent measurement of whether potentially reversible causes of infertility were evaluated or treated before progression to IVF (**Table 1** and **Table 2**). Interventions were identified using prespecified CPT and HCPCS procedure codes, ICD-10-CM diagnosis and encounter codes when applicable, and National Drug Codes for pharmacy-dispensed medications. For each cohort, cumulative proportions of patients receiving the relevant guideline-recommended service were calculated at defined intervals following the index infertility diagnosis, and these proportions were then compared with the cumulative proportion who had initiated IVF during the same follow-up windows.

**Table 1:** Female Infertility Recommendations Evaluated in the Study.

| Condition | Practice Recommendation | Supporting Quote | Clinical Rationale |
| --- | --- | --- | --- |
| **Amenorrhea** | Hormonal evaluation | “Core elements ...FSH, LH, AMH, estradiol...”<sup>6</sup> p.56 | Amenorrhea is a symptom requiring etiologic diagnosis before fertility treatment decisions are made. |
| **Amenorrhea** | Pelvic ultrasound | “Pelvic ultrasound can be highly informative and, if accessible and available, should be considered at the forefront of investigations for the evaluation of secondary amenorrhea.”<sup>6</sup> p.58 | Ultrasound may identify structural, ovarian, and endocrine contributors to infertility. |
| **Dysmenorrhea / Suspected Endometriosis** | Laparoscopy | “...a surgical procedure such as a laparoscopy is required for definitive diagnosis of endometriosis... significant findings suggestive of endometriosis [include] cyclic or chronic pelvic pain, dysmenorrhea.”<sup>7</sup> p.592 | Surgical evaluation may identify and treat endometriosis contributing to infertility. |
| **Endometriosis (<35 years)** | Expectant management / SO-IUI | “In younger women (under age 35 years) with stage I/II endometriosis-associated infertility, expectant management or SO/IUI can be considered as first-line therapy.”<sup>7</sup> p.596 | Less invasive treatment options may achieve pregnancy before IVF is required. |
| **Hyperprolactinemia** | Dopamine agonists | “In the absence of another organic condition, dopamine agonists are the preferred treatment of hyperprolactinemia...”<sup>6</sup> p.53 | Hyperprolactinemia is often a reversible endocrine cause of infertility. |
| **Hypothyroidism** | Thyroid evaluation | “Abnormal TSH values should be followed up by a repeat value in addition to serum thyroxine (tetraiodothyronine; T4 or free T4) levels and treated accordingly”<sup>6</sup> p.53 | Thyroid dysfunction may contribute to infertility, ovulatory dysfunction, and miscarriage risk. |
| **Hypothyroidism** | Thyroid treatment | “Levothyroxine should remain the standard of care for treating hypothyroidism”<sup>18</sup> p.1670 | Overt hypothyroidism can potentially have a significant impact on reproductive outcomes. |
| **Polycystic Ovary Syndrome (PCOS)** | Gonadotropins | “Gonadotrophins alone could be considered rather than clomiphene citrate...”<sup>5</sup> p.785 | Gonadotropins represent an alternative ovulation-induction strategy prior to IVF. |
| **PCOS** | Letrozole | “Letrozole should be the first-line pharmacological treatment for ovulation induction.”<sup>5</sup> p.784 | Letrozole is recommended as first-line ovulation induction therapy for anovulatory infertility associated with PCOS. |
| **PCOS** | Metformin | “Metformin could be used alone, in women with PCOS with anovulatory infertility.”<sup>5</sup> p.785 | Metformin may improve ovulation and address metabolic contributors to infertility. |
| **PCOS** | Ovarian surgery | “Laparoscopic ovarian surgery could be second-line therapy.”<sup>5</sup> p.786 | Surgical restoration of ovulation may reduce the need for IVF in selected patients. |

**Table 2.** Male Infertility Recommendations Evaluated in the Study.

| **Condition** | **Practice Recommendation** | **Supporting Quote** | **Clinical Rationale** |
| --- | --- | --- | --- |
| **Azoospermia** | Genetic testing | “Karyotype and Y-chromosome microdeletion analysis...”<sup>8</sup> p.55 | Genetic testing may identify underlying chromosomal or genetic causes of male infertility. |
| **Azoospermia** | Hormonal evaluation | “Clinicians should obtain hormonal evaluation including follicle-stimulating hormone and testosterone for infertile males...”<sup>8</sup> p.55 | Hormonal testing assists in distinguishing impaired sperm production from obstructive causes of azoospermia. |
| **Azoospermia** | Test for Cystic Fibrosis Transmembrane Conductance Regulator (CFTR) gene | “CFTR mutation...testing...in men with vasal agenesis or idiopathic obstructive azoospermia”<sup>8</sup> p.55 | CFTR testing may identify cystic fibrosis–related reproductive tract abnormalities associated with obstructive azoospermia. |
| **Male infertility** | Repeat semen analysis | “...at least two SAs...are important to obtain...”<sup>8</sup> p.54 | Repeated testing accounts for biologic variability in semen parameters and improves diagnostic accuracy. |
| **Male Infertility** | Semen analysis | “Initial evaluation of the male should also include one or more semen analyses (SAs).”<sup>8</sup> p.54 | Semen analysis is the foundational diagnostic test in the evaluation of male infertility. |
| **Recurrent Pregnancy Loss** | Karyotype and Sperm DNA fragmentation | “For couples with RPL, men should be evaluated with karyotype and sperm DNA fragmentation.”<sup>8</sup> p.58 | Male genetic abnormalities and sperm DNA damage may contribute to recurrent pregnancy loss. |
| **Varicocele** | Varicocelectomy | “Surgical varicocelectomy should be considered in men attempting to conceive, who have palpable varicocele(s), infertility, and abnormal semen parameters...”<sup>9</sup> p.62 | Varicocele repair may improve semen parameters and increase the likelihood of conception. |

### Guideline Measures

## ANALYSIS

We calculated cumulative adherence and cumulative IVF initiation rates reported at 1 month, 3 months, 6 months, and 9 months following the initial infertility diagnosis. For each recommendation, cumulative guideline adherence was defined as the proportion of eligible patients who had undergone the recommended diagnostic evaluation or treatment by a specified time point, while cumulative IVF utilization was defined as the proportion of patients who had initiated IVF during the same interval. Consistent with established approaches in quality improvement, implementation science, and guideline adherence research, we calculated care gaps as absolute percentage-point between recommendations and observed clinical practice.^19^

A care gap refers to the difference between guideline-recommended care and the actual care that is delivered to patients over a specified time period. In healthcare quality measurement, care gaps are used to identify instances in which recommended investigations, treatments, preventive services, or follow-up interventions have not occurred despite a clinical indication.^20^ Care-gap analysis has become a common quality-improvement methodology because it quantifies the extent to which real-world practice deviates from established standards of care and helps identify opportunities for improving patient outcomes. The concept is closely related to the broader quality literature describing gaps between recommended and actual healthcare delivery.^21–22^

For our analysis, an IVF–Guideline Care Gap metric was calculated as the difference between cumulative IVF utilization and cumulative guideline adherence at each time point

### Care Gap = IVF Utilization [%] - Guideline Adherence [%]

A positive care gap indicates that a recommended intervention was not completed prior to IVF for a proportion of patients, whereas a negative care gap suggests that care was completed before progression to IVF (recall, that by design, all patients in our analysis proceeded to IVF). Quantifying care gaps allows investigators to move beyond simple reporting of utilization rates and instead evaluate adherence to clinical guidelines, identify areas of underuse or delayed care, and assess opportunities for quality improvement.

The magnitude of the care gap therefore serves as a measure of the implementation gap between published recommendations and real-world clinical practice. Similar approaches have been widely employed in health-services research to evaluate treatment gaps, evidence-to-practice gaps, and quality-of-care variation across medical specialties.^23^

## RESULTS

### One Month Guideline Adherence Versus IVF Initiation

By one month after diagnosis, between 4.7% and 12.3% of patients had already initiated IVF depending on the underlying diagnosis, whereas adherence to many guideline-recommended evaluations and treatments remained low. For example, only 1.0% of men had completed the recommended two semen analyses, 0.6% of men with paternity of recurrent pregnancy loss had undergone sperm DNA fragmentation testing, 0.8% of women younger than 35 years with endometriosis had undergone IUI, and 0.3% of women with PCOS had undergone laparoscopic ovarian surgery. These findings demonstrate that IVF initiation frequently began before completion of several recommended diagnostic and therapeutic interventions **(Table 3)**.

**Table 3:** Adherence versus IVF initiation at one month (female and male conditions)

| Clinical condition (as determined by ICD10 codes submitted for claim) N97.X for female, N46.X for male | Practice Recommendation | N* | Month 1 Adherence (%) | Month 1 IVF (%) |
| --- | --- | --- | --- | --- |
| Amenorrhea | Human chorionic gonadotropin (pregnancy test) | 899 | 67.3 | 4.7 |
| Amenorrhea | Pelvic Ultrasound | 899 | 34.7 | 4.7 |
| Amenorrhea | Serum AMH | 899 | 3.8 | 4.7 |
| Amenorrhea | Serum Estradiol | 899 | 30.3 | 4.7 |
| Amenorrhea | Serum FSH | 899 | 16.0 | 4.7 |
| Amenorrhea | Serum LH | 899 | 18.0 | 4.7 |
| Dysmenorrhea | Laparoscopy | 1002 | 4.2 | 5.0 |
| Endometriosis | Intrauterine Insemination | 430 | 1.4 | 7.0 |
| Hyperprolactinemia | Bromocriptine | 482 | 3.7 | 5.4 |
| Hyperprolactinemia | Cabergoline | 482 | 9.1 | 5.4 |
| Hypothyroidism | Levothyroxine | 1801 | 24.9 | 8.4 |
| Oligomenorrhea or Androgen Excess – 17-OHP | Serum 17-hydroxyprogesterone | 3452 | 5.8 | 8.9 |
| Polycystic Ovary Syndrome (PCOS) | Gonadotropins | 2201 | 4.5 | 12.3 |
| PCOS | Letrozole | 2201 | 8.1 | 12.3 |
| PCOS | Metformin | 2201 | 7.1 | 12.3 |
| PCOS | Ovarian Surgery | 2201 | 0.3 | 12.3 |
| Azoospermia | Karyotype and Y-Microdeletion | 52 | 17.3 | 7.7 |
| Azoospermia | Serum FSH | 52 | 26.9 | 7.7 |
| Azoospermia | Serum Testosterone | 52 | 28.8 | 7.7 |
| Azoospermia | Test for Cystic Fibrosis Transmembrane Conductance Regulator gene | 52 | 3.8 | 7.7 |
| Male Infertility | ≥2 Semen Analyses | 485 | 1.0 | 10.3 |
| Recurrent Pregnancy Loss | Karyotype | 346 | 11.8 | 6.4 |
| Recurrent Pregnancy Loss | Sperm DNA Fragmentation | 346 | 0.6 | 6.4 |
| Varicocele | Varicocelectomy | 87 | 2.3 | 8.0 |

\*(Female infertility Total N= 9884; Male infertility Total N=485)

### Female Guideline Adherence Versus IVF Initiation

Across female infertility conditions, adherence to guideline-recommended evaluations and treatments increased over time but generally remained below IVF initiation rates. By 3 months, IVF initiation ranged from 28% to 39% (varying by underlying diagnosis), while adherence to many recommended interventions remained below 20%. By 6 months, IVF initiation had increased to 52%–71%, whereas adherence to many treatments and diagnostic evaluations remained substantially lower, with only 4 of 16 interventions having adherence >50%. By 9 months, IVF initiation ranged from 70% to 86%, yet adherence remained below 40% for many condition-specific therapies, including medication use in PCOS (PMOS) - letrozole (34.1%), metformin (22.1%), gonadotropins (35.3%); medication in hyperprolactinemia, cabergoline (25.7%), IUI for endometriosis (9.3%), and laparoscopy for dysmenorrhea (11.6%). In contrast, some diagnostic evaluations such as pelvic ultrasound, estradiol testing, and hCG testing exceeded IVF initiation rates. (**Table 4**).

**Table 4:**
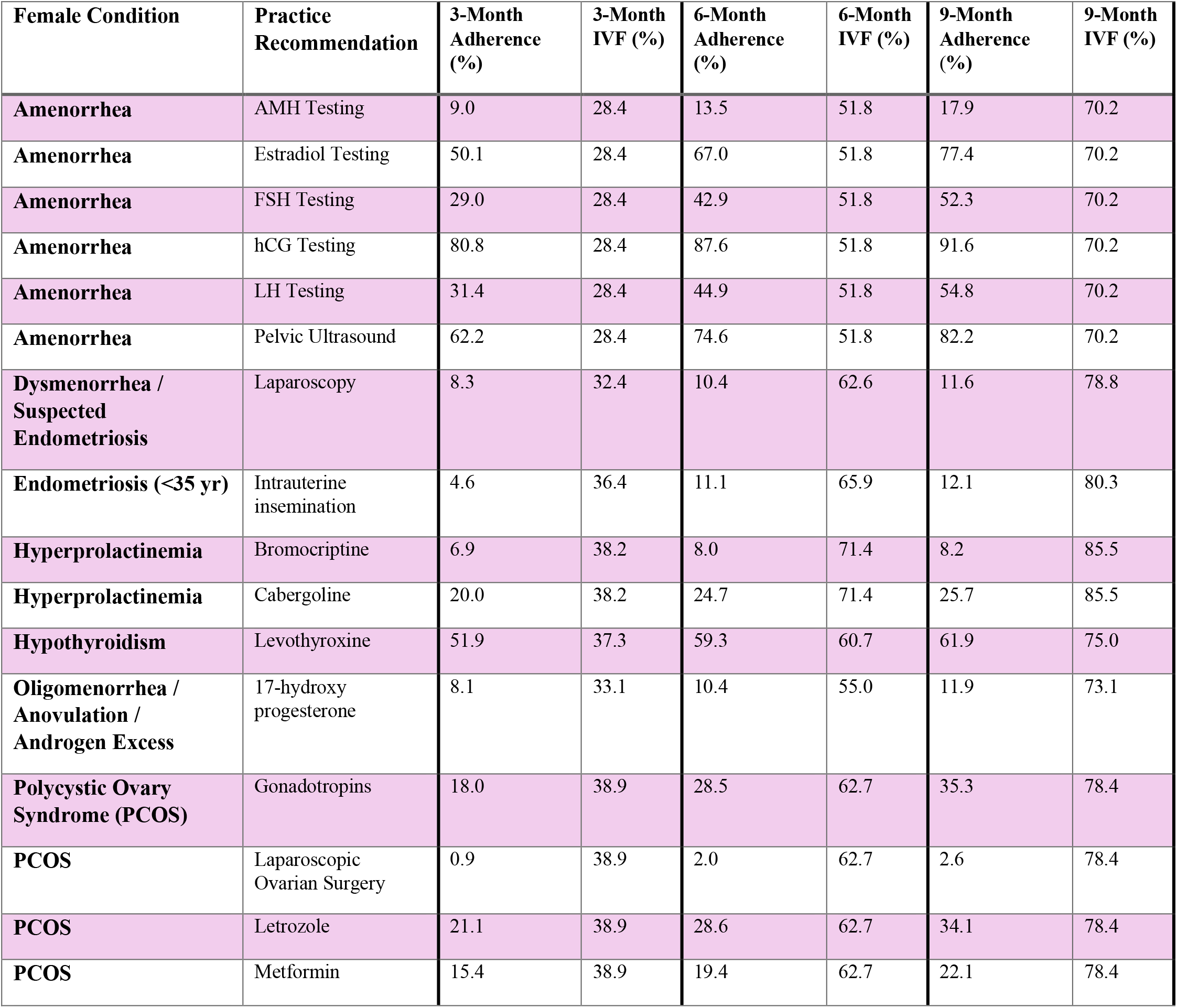
Adherence versus IVF Initiation over 9 Months (Female Conditions)

### Male Guideline Adherence Versus IVF Initiation

A similar pattern was observed among male infertility recommendations. At 3 months, IVF initiation ranged from 33% to 38% (varying by underlying diagnosis), while adherence to several recommended male evaluations remained low, including completion of two semen analyses (6.0%), varicocelectomy for diagnosed varicocele (9.2%), karyotype and y-chromosome microdeletion for azoospermia (19.2%), and sperm DNA fragmentation testing for recurrent pregnancy loss (1.5%). By 6 months, IVF initiation had increased to 54%–66%, whereas adherence remained relatively unchanged for many recommendations. By 9 months, IVF initiation ranged from 73% to 81%, while adherence remained 7.8% for two semen analyses, 14.5% for varicocelectomy for diagnosed varicocele, 20.8% for karyotype and y-chromosome microdeletion, and so on. Although adherence generally increased over time, the proportion with IVF initiation consistently exceeded completion of many guideline-recommended male infertility evaluations and treatments throughout follow-up (**Table 5**).

**Table 5:**
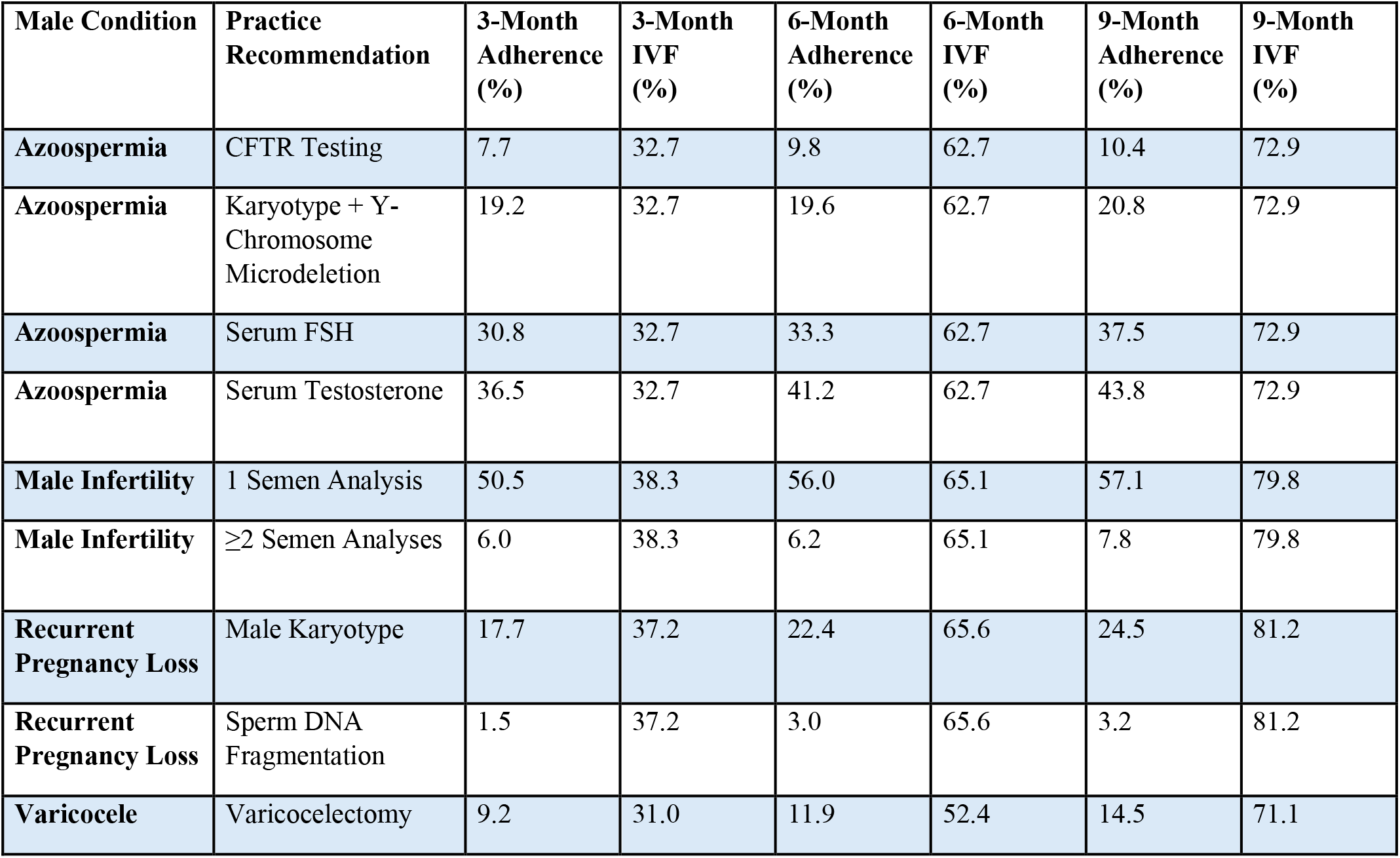
Adherence versus IVF Over 9 Months (Male Conditions)

| **Male Condition** | **Practice Recommendation** | **3-Month Adherence (%)** | **3-Month IVF (%)** | **6-Month Adherence (%)** | **6-Month IVF (%)** | **9-Month Adherence (%)** | **9-Month IVF (%)** |
| --- | --- | --- | --- | --- | --- | --- | --- |
| **Azoospermia** | CFTR Testing | 7.7 | 32.7 | 9.8 | 62.7 | 10.4 | 72.9 |
| **Azoospermia** | Karyotype + Y-Chromosome Microdeletion | 19.2 | 32.7 | 19.6 | 62.7 | 20.8 | 72.9 |
| **Azoospermia** | Serum FSH | 30.8 | 32.7 | 33.3 | 62.7 | 37.5 | 72.9 |
| **Azoospermia** | Serum Testosterone | 36.5 | 32.7 | 41.2 | 62.7 | 43.8 | 72.9 |
| **Male Infertility** | 1 Semen Analysis | 50.5 | 38.3 | 56.0 | 65.1 | 57.1 | 79.8 |
| **Male Infertility** | ≥2 Semen Analyses | 6.0 | 38.3 | 6.2 | 65.1 | 7.8 | 79.8 |
| **Recurrent Pregnancy Loss** | Male Karyotype | 17.7 | 37.2 | 22.4 | 65.6 | 24.5 | 81.2 |
| **Recurrent Pregnancy Loss** | Sperm DNA Fragmentation | 1.5 | 37.2 | 3.0 | 65.6 | 3.2 | 81.2 |
| **Varicocele** | Varicocelectomy | 9.2 | 31.0 | 11.9 | 52.4 | 14.5 | 71.1 |

### Care Gap Analysis

Among female infertility cohorts, the largest treatment care gaps were observed for bromocriptine treatment of hyperprolactinemia (77.3%), laparoscopic ovarian surgery for PCOS (75.8%), intrauterine insemination among women younger than 35 years with endometriosis (71%), and laparoscopy among women with dysmenorrhea suggestive of endometriosis (67%). Adherence to diagnostic evaluation for amenorrhea was generally higher, with pelvic ultrasound, estradiol testing, and hCG testing exceeding IVF initiation rates by month 9; however, these treatments may have been done as part of the IVF process itself (**Table 6**).

**Table 6:** Female Guideline Care Gaps (Ranked by Highest Care Gap)

| **Female Condition** | **Practice Recommendations** | **Adherence at 9 Months (%)** | **IVF Initiated at 9 Months (%)** | **Absolute Care Gap (%) \*** |
| --- | --- | --- | --- | --- |
| **Hyperprolactinemia** | Bromocriptine | 8.2 | 85.5 | **77.3** |
| **PCOS** | Laparoscopic Ovarian Surgery | 2.6 | 78.4 | **75.8** |
| **Endometriosis (<35 Years)** | Intrauterine Insemination (IUI) | 9.3 | 80.3 | **71.0** |
| **Dysmenorrhea / Suspected Endometriosis** | Laparoscopy | 11.6 | 78.8 | **67.2** |
| **Hyperprolactinemia** | Cabergoline | 25.7 | 85.5 | **59.8** |
| **PCOS** | Metformin | 22.1 | 78.4 | **56.3** |
| **Amenorrhea** | Anti-Müllerian Hormone (AMH) Testing | 17.9 | 70.2 | **52.3** |
| **PCOS** | Letrozole | 34.1 | 78.4 | **44.3** |
| **PCOS** | Gonadotropins | 35.3 | 78.4 | **43.1** |
| **Amenorrhea** | Follicle-Stimulating Hormone (FSH) Testing | 52.3 | 70.2 | **17.9** |
| **Amenorrhea** | Luteinizing Hormone (LH) Testing | 54.8 | 70.2 | **15.4** |
| **Hypothyroidism** | Levothyroxine | 61.9 | 75.0 | **13.1** |
| **Amenorrhea** | Estradiol Testing | 77.4 | 70.2 | **-7.2** |
| **Amenorrhea** | Pelvic Ultrasound | 82.2 | 70.2 | **-12.0** |
| **Amenorrhea** | Human Chorionic Gonadotropin (hCG) Testing | 91.6 | 70.2 | **-21.4** |

\*Absolute Care Gap (%) = IVF initiation rate – guideline adherence rate. Positive values indicate that the proportion with IVF initiation exceeded the proportion completing of the guideline-recommended evaluation or treatment. Negative values indicate the recommended evaluation, or treatment was performed more frequently than IVF initiation.

By 9 months following infertility diagnosis, the largest care gaps were observed for sperm DNA fragmentation testing in recurrent pregnancy loss (78.0%), completion of two semen analyses (70.5% and testing for the Cystic Fibrosis Transmembrane Conductance Regulator (CFTR) gene among men with azoospermia (62.5%) (**Table 7**).

**Table 7:** Male Guideline Care Gaps (Ranked by Highest Care Gap)

| Male Condition | Practice Recommendations | Adherence at 9 Months (%) | IVF Initiated at 9 Months (%) | Absolute Care Gap (%) * |
| --- | --- | --- | --- | --- |
| **Recurrent Pregnancy Loss (male partner)** | DNA fragmentation testing | 3.2 | 81.2 | **78.0** |
| **Male infertility** | ≥2 semen analyses | 9.3 | 79.8 | **70.5** |
| **Azoospermia** | CFTR testing | 10.4 | 72.9 | **62.5** |
| **Recurrent Pregnancy Loss (male partner)** | Karyotype testing | 24.5 | 81.2 | **56.7** |
| **Varicocele** | Varicocelectomy | 14.5 | 71.1 | **56.6** |
| **Azoospermia** | Genetic testing (karyotype + Y-chromosome microdeletion) | 20.8 | 72.9 | **52.1** |
| **Azoospermia** | FSH testing | 37.5 | 72.9 | **35.4** |
| **Azoospermia** | Testosterone testing | 43.8 | 72.9 | **29.1** |
| **Male infertility** | 1 semen analysis | 57.1 | 79.8 | **22.7** |

\*Absolute Care Gap (%) = IVF initiation rate – guideline adherence rate. Positive values indicate that the proportion with IVF initiation exceeded the proportion completing of the guideline-recommended evaluation or treatment. Negative values indicate the recommended evaluation, or treatment was performed more frequently than IVF initiation.

## DISCUSSION

This analysis identified substantial gaps between published infertility recommendations and observed clinical practice among commercially insured patients who ultimately underwent IVF. These gaps were evident across both female- and male-factor infertility pathways, although the specific patterns differed by diagnosis. The divergence between IVF utilization and guideline adherence was already apparent in the first month, widened further by 6 months, and remained substantial at 9 months for many recommendations in both female and male infertility populations. Viewed in light of the conceptual distinction between IVF and restorative reproductive medicine (RRM), the findings suggest that many patients may proceed to a treatment pathway that bypasses reproductive dysfunction before there is documented evidence that potentially reversible contributors have been fully evaluated, corrected, or given sufficient time to respond to treatment.

### Diagnosis-Specific Care Gaps

Several patterns emerged. First, recommended diagnostic evaluations appeared underutilized. Male infertility evaluation showed particularly low adherence to the recommendation to complete two semen analyses, despite the recognized biological variability of semen parameters. Similarly, endocrine and genetic evaluation among men with azoospermia occurred in only a minority of patients. These findings are clinically important because male-factor infertility may reflect endocrine, genetic, anatomic, or potentially correctable contributors that require diagnosis before management decisions can be appropriately individualized. Infertility may also be sign of serious undiagnosed medical conditions that left untreated may have a negative impact on the patient’s long-term health or contribute to earlier mortality.^24^

Second, potentially reversible causes of infertility frequently appeared undertreated before IVF. Among women with PCOS (PMOS), relatively low rates of first-line ovulation induction therapy, insulin-sensitizing therapy, and second-line treatment options were documented before IVF became widely utilized. Similar findings were observed among patients with hyperprolactinemia and thyroid disorders. These conditions are not merely labels associated with infertility; they may represent modifiable endocrine or metabolic dysfunction that can impair ovulation, endometrial receptivity, pregnancy maintenance, and overall health and well-being.^25–26^

Third, surgical evaluation and treatment appeared uncommon in cohorts for whom professional guidelines support consideration of operative management. This pattern was evident among male infertility patients with varicocele and among female patients with endometriosis or dysmenorrhea. In an RRM framework, these findings are particularly important because preservation and correction of reproductive anatomy are central components of care. When clinically indicated surgery is not considered IVF, an opportunity to correct underlying anatomic contributors may be missed.

Taken together, these female- and male-factor care gaps suggest a common implication: potentially reversible or clinically informative causes of infertility may be missed, incompletely evaluated, or insufficiently treated before IVF. In women, endocrine, metabolic, inflammatory, and anatomic abnormalities such as PCOS (now polyendocrine ovarian metabolic disorder, PMOS), hyperprolactinemia, hypothyroidism, and endometriosis may impair fertility but remain amenable to diagnosis-directed treatment. In men, repeat semen analysis, endocrine testing, genetic evaluation, and treatment of varicocele may identify contributors that alter prognosis, management, counseling, or treatment choice.

The timing of care is also important. Correction of reproductive dysfunction is not instantaneous. Hormonal normalization, improved ovulation, metabolic response, recovery after surgery, and improvement in spermatogenesis or semen parameters may require time, follow-up, and treatment adjustment before any fertility benefit becomes apparent. Progression to IVF before abnormalities are identified, corrected, and given time to respond may narrow the opportunity for less invasive, restorative care.

### Comparison With Broader Care-Gap Literature

The magnitude of the care gaps observed in this analysis appears clinically meaningful. In broader quality-of-care literature, incomplete adherence to evidence-based recommendations is common;^27^ however, the degree of underuse observed here is especially important because the missed steps involve diagnostic evaluation and treatment of potentially reversible causes of infertility before escalation to IVF. These findings support the need for initial evaluation and treatment according to RRM principles.

These findings are consistent with the wider quality-improvement literature, which has repeatedly shown gaps between recommended care and care delivered.^28^ In the infertility context, however, these gaps have a distinctive implication. Low rates of repeat semen analysis, endocrine and genetic evaluation in azoospermia, ovulation induction in PCOS, treatment of endocrine disorders, and consideration of indicated reproductive surgery should not be interpreted as minor documentation issues alone. Rather, they suggest that many patients may proceed to IVF before completion of guideline-supported evaluation or diagnosis-directed treatment. From an RRM perspective, the issue is not whether IVF can achieve pregnancy, but whether patients receive a meaningful opportunity for diagnosis, correction, restoration, and addressing long-term health before reproductive function is bypassed.

### Interpretation and Implications

Although claims data cannot determine clinical reasoning, patient preferences, prior evaluations, contraindications, or services paid for outside insurance, the magnitude and consistency of the observed gaps suggest that many patients may proceed to IVF without documented completion of the diagnostic and therapeutic steps recommended by ASRM and AUA/ASRM.

The gaps identified do not diminish the role of IVF for patients who choose, or benefit from it. Rather, they highlight a quality-of-care concern in the pathway leading to IVF.

RRM is directly relevant to these gaps because its clinical focus is the systematic identification, correction, restoration, and preservation of reproductive function and anatomy. RRM is designed to ensure that both partners receive full evaluation, that underlying causes are actively sought, and that treatable contributors are addressed.^1,29^ In this sense, RRM is not simply an alternative to IVF; it is a quality-of-care framework directed at the gaps identified in this study. Addressing these gaps may improve patient-centered care, better align treatment with published recommendations, and ensure that couples are not moved prematurely into IVF when potentially reversible contributors remain unaddressed.^30^

There are also implications for healthcare expenditures. Many recommended evaluations and treatments are substantially less expensive than IVF. Future analyses should evaluate whether improved adherence to infertility guidelines affects IVF utilization patterns, total costs of care (including not just the IVF service itself but the comprehensive prenatal, birth and postpartum costs for mother and baby), live birth rates, time to pregnancy, obstetric outcomes (maternal and fetal), patient satisfaction, and patient-reported experience of care.

## STRENGTHS AND LIMITATIONS

This study has several important strengths. It draws on a large national commercial claims database, providing a broad real-world view of infertility-care patterns across a substantial insured population. The longitudinal design allowed tracking of care over time from initial infertility diagnosis to IVF utilization, making it possible to examine the sequencing of evaluation and treatment rather than isolated interventions. In addition, the analysis spans multiple infertility diagnoses and guidelines.

This study also has important limitations. Because the analysis relies on administrative claims, it reflects billed services rather than clinical intent or the full context of decision-making. Treatments, procedures, or evaluations paid for outside insurance may not be captured, and eligibility for specific guideline-recommended interventions cannot always be determined from claims alone. Key clinical details, including disease severity, patient preferences, prior workup (outside the 15 months window or insurance coverage), contraindications, and clinician rationale, were unavailable. Therefore, nonreceipt of a service cannot always be interpreted as underuse, and in some cases may reflect appropriate individualized care. In addition, the observed associations are descriptive and cannot establish causation. Females and males were analyzed separately, because the data structure did not allow couples-level analysis. In at least one case, the recommendations are “overlapping,” e.g., hyperprolactinemia would be appropriately treated either by bromocriptine or cabergoline, not both medications at the same time. Some recommendations would be considered secondary, not primary, e.g., laparoscopic ovarian surgery for PCOS. We did not assess the level of evidence for each recommendation, and we did not assess the level of acceptance for each recommendation among practicing clinicians.

Nevertheless, these limitations do not negate the importance of the observed patterns. Even allowing for incomplete claims capture and individualized clinical decision-making, the low documented rates of multiple recommended diagnostic and treatment steps suggest that recommended evaluation and intervention steps are often not completed before initiating IVF. This deserves further evaluation in more robust data sets, such as data based on electronic health records (not just claims data).

## CONCLUSIONS

Among commercially insured infertility patients who subsequently underwent IVF, adherence to multiple ASRM and AUA/ASRM diagnostic and treatment recommendations appeared to be limited. Many testing, medical treatment, and surgical management recommendations were completed in only a minority of patients before IVF initiation across several diagnosis-specific groups.

These findings raise important questions about whether patients receive a meaningful opportunity for comprehensive diagnosis-directed care before proceeding to IVF.^31^ From an RRM perspective, the observed gaps are clinically significant because they suggest that potentially reversible causes of infertility may remain unidentified or insufficiently treated before reproductive function is bypassed through assisted reproduction, potentially compromising the patients’ long-term health and possibly introducing unnecessary risks for pregnancy and live birth.

Further research is needed to evaluate implementation of infertility guidelines, identify barriers to comprehensive pre-IVF evaluation and treatment, and determine whether restorative, diagnosis-directed models of care can improve patient-centered outcomes, reduce unnecessary escalation to IVF, and better align infertility care with appropriate professional recommendations and patient preferences.

## ETHICS STATEMENT

This study utilized de-identified administrative claims data. No direct patient contact occurred, and no identifiable personal health information was accessed by the investigators.

## AUTHOR CONTRIBUTIONS

**TAP:** Conceptualization, Study design, Data acquisition and analysis, Interpretation of findings, Manuscript drafting, Writing - Review & Editing. **TP:** Conceptualization, Study design, Data acquisition and analysis, Interpretation of findings, Writing - Review & Editing. **MM:** Study design, Interpretation of findings, Writing - Review & Editing. **CT:** Study design, Interpretation of findings, Writing - Review & Editing. All authors read and approved the final manuscript.

## DATA AVAILABILITY STATEMENT

The MarketScan® Commercial Claims and Encounter Database used for this study is proprietary and subject to licensing restrictions. Data may be available from the data owner subject to applicable agreements and permissions.

